# Diagnostic accuracy of anti-HlyE and LPS antibodies in typhoid intestinal perforation in Niger

**DOI:** 10.64898/2026.10.06.26364924

**Authors:** Ezekiel B. Ackah, Beatrice Nyantakyiwaa, Michael Owusu, Isaac Kissi, Katherine Shafer, Leah Sukri, Laura Hobbs, Issoufou Samaila, Shrimati Datta, Claire Munroe, Yaw Adu Sarkodie, Ellis Owusu-Dabo, Jason R. Andrews, Kathleen M. Neuzil, Yakoubou Sanoussi, Matthew B. Laurens, Richelle C. Charles

## Abstract

Typhoid intestinal perforation (TIP) is a late complication of *Salmonella enterica* serovar Typhi infection, and blood cultures, if available, are often negative at this stage, making etiologic confirmation challenging. In areas without access to blood culture diagnostics, TIP incidence can serve as a proxy for enteric fever burden. Antibodies against hemolysin E (HlyE) and *S*. Typhi lipopolysaccharide (LPS) are promising serologic markers for typhoid, but their diagnostic utility for TIP remains unclear. In this study, we assessed the diagnostic accuracy of anti-HlyE and anti-LPS IgG and IgA antibodies measured by kinetic enzyme immunoassay in children undergoing emergency laparotomy for acute abdomen in Niger. Dried blood spots (DBS) were collected pre-operatively (baseline) and within 30 days post-hospital discharge (convalescence). Patients classified intraoperatively as TIP (*n* = 341) or as confirmed non-typhoid-related alternative etiology (i.e., controls, *n* = 48) were included in this serologic analysis. The primary analyses were conducted in a longitudinal sub-cohort of 122 TIP cases and 33 controls in whom all four anti-HlyE and anti-LPS IgA and IgG responses were measured at baseline and at convalescence. Anti-HlyE IgA and anti-LPS IgA were additionally measured at baseline in the full analytic cohort. In the primary analysis, all antigen-antibody isotype combinations distinguished TIP cases from controls with high accuracy: anti-HlyE IgA (AUC 0.94), anti-HlyE IgG (AUC 0.94), anti-LPS IgA (AUC 0.96), and anti-LPS IgG (AUC 0.96). Anti-HlyE IgA and anti-LPS IgA performed similarly in the full cohort (AUC 0.93 and 0.93, respectively), supporting the representativeness of the sub-cohort. Anti-LPS IgG showed the best overall performance (99% sensitivity, 94% specificity), exceeding that of all other individual markers, and combining markers did not improve diagnostic accuracy. Diagnostic performance at convalescence was similar to or lower than that observed at presentation. Overall, anti-HlyE and anti-LPS antibodies showed high diagnostic accuracy for TIP at presentation, extending the potential utility of these serologic markers for supporting the diagnosis of typhoid-associated intestinal perforation and improving surveillance of severe typhoid disease when patients present with late-stage complications. Our findings show that serologic testing could improve case detection and burden estimation of enteric fever and its complications in resource-limited settings.

## INTRODUCTION

Typhoid fever is a severe systemic infection caused by *Salmonella enterica* serovar Typhi (*S*. Typhi), a human-restricted pathogen [1]. The Global Burden of Disease Study estimated that, in 2023, there were 6.2 million new cases of typhoid fever globally, resulting in ∼72,000 deaths in that year [2]. South Asia accounted for 73% of the global cases, with an estimated incidence of 237 per 100,000 person-years, while sub-Saharan Africa (SSA) accounted for 14% of cases, with an estimated incidence of 73 per 100,000 person-years [2]. Despite South Asia reporting most cases of typhoid fever and mortality, the SSA region had a higher case-fatality rate (CFR), estimated at 1.4% in 2023 as opposed to 1.1% CFR in South Asia [2]. This outcome likely reflects the limited access to healthcare in the SSA region, including delays in diagnosis, initiation of appropriate treatment and management of complications, as well as increasing prevalence and spread of multidrug-resistant *S.* Typhi in the region [3–5]. Recent community-based serosurveillance studies suggest that the true burden of typhoid infection is substantially underestimated by blood culture-based surveillance, with seroincidence estimates up to 16–32 times higher than reported clinical incidence, indicating a large burden of unrecognized infections in endemic settings [6–8].

Delayed diagnosis and emergence of multidrug resistance increase the risk of disease progression and severe complications, including typhoid intestinal perforation (TIP), which remains common in endemic settings such as Niger, especially among children [9–12]. TIP typically occurs approximately two weeks after initial symptoms. It is caused by prolonged bacterial invasion of Peyer’s patches in the terminal ileum, leading to inflammation, necrosis, and ulceration that progressively extends through the intestinal wall, ultimately causing full-thickness perforation and peritonitis [13, 14]. Despite the substantial burden and severity of TIP in Africa, laboratory confirmation of *S.* Typhi remains challenging as blood cultures have both limited availability and limited sensitivity, particularly in patients with intestinal perforation [4, 12, 15, 16]. Consequently, clinically and intraoperatively diagnosed cases of intestinal perforation are often not microbiologically confirmed [15].

Anti-HlyE and anti-LPS antibodies show ∼90% sensitivity and specificity for diagnosing acute enteric fever [17–20]. Their accuracy in diagnosing TIP has not yet been characterized. These biomarkers could serve as reliable and accessible diagnostic tools to estimate the incidence of TIP-associated typhoid disease in resource-limited areas. In this study, we aimed to evaluate the ability of the anti-HlyE and anti-LPS antibodies in identifying patients with surgically confirmed TIP compared with those with other confirmed causes of acute abdomen for emergency laparotomy.

## MATERIALS AND METHODS

### Ethical Statement

Ethical approval was obtained from the institutional review boards and/or the research and ethical review committees in Niger (Hôpital de la SIM-Galmi, N°64/2023/CNERS) and the United States of America (University of Maryland, Baltimore, HP-00103732; and Mass General Brigham, 2019P000152). The study was carried out in accordance with the ethical principles outlined in the Nuremberg Code, the Belmont Report, and the Declaration of Helsinki [21]. Before participation, written informed consent was obtained from the legal guardians of all participants, in addition to assent from participants aged 15–17 years.

### Study Population, Specimen Collection, and Selection of Participants for Serologic Analysis

This analysis leveraged samples collected from a hospital-based prospective study of typhoid intestinal perforation in Galmi, a rural settlement located in the Tahoua Region of southern Niger [22]. Patients aged 3–17 years were enrolled from August 2023 to December 2024 if they presented to or were referred to Hôpital de la SIM with a clinical diagnosis of an atraumatic acute abdomen requiring emergency laparotomy. Whole blood samples were collected into BACT/ALERT^®^ bottles (bioMérieux, Marcy l’Étoile, France) for blood culture to isolate *S*. Typhi as previously described [23], and on TropBioTM filter papers (Cellabs, Brookvale, NSW, Australia) for dried blood spots (DBS) at presentation (baseline or acute stage); DBS samples were also collected within 30 days post-hospital discharge (convalescence). DBS were stored with desiccant at-20°C or lower and shipped at room temperature to the Kwame Nkrumah University of Science and Technology (KNUST) in Kumasi, Ghana, for anti-HlyE and anti-LPS antibody testing.

All participants enrolled in the parent study who underwent surgical intervention or had an available dried blood spot (DBS) sample were eligible for inclusion in this analysis. Participants with suspected typhoid-related disease but without confirmed intestinal perforation were excluded to reduce outcome misclassification in the primary diagnostic accuracy analysis. Inclusion of these participants would likely bias estimates toward reduced discrimination because some may represent true typhoid disease without pathognomonic perforation. The full analysis cohort consisted of all remaining participants, and baseline anti-HlyE and anti-LPS IgA were measured in these individuals. A nested longitudinal sub-cohort (primary analysis cohort) included participants with paired acute and convalescent samples, with anti-HlyE and anti-LPS IgA and IgG responses measured at both time points. All eligible controls were included in his analysis and TIP cases were randomly selected for this sub-cohort at a ratio of 4 cases to 1 control, while maintaining representation across all age groups (< 5, 5–10, 11–15, and > 15 years).

### Sample Size

A minimum sample size of 145 participants was estimated to assess the diagnostic performance of the HlyE and LPS antibodies in the assay at baseline and convalescence. This was calculated assuming 90% sensitivity and specificity, 10% marginal error, and 5% (*α* = 0.05) type I error [24], using a TIP prevalence of 70% among cases of peritonitis in the Tahoua Region in southern Niger [13].

### Immunoassays for Anti-HlyE and Anti-LPS Antibodies

The DBS samples were eluted and tested for IgA and IgG antibodies against *S*. Typhi HlyE and LPS antigens using a kinetic enzyme-linked immunosorbent assay (ELISA), as previously described [6]. One DBS ear was eluted overnight at 4°C in 66.7 µL phosphate-buffered saline (PBS) with 0.05% Tween-20. ELISA plates were coated with HlyE (1 µg/mL) or LPS (2.5 µg/mL) antigens and incubated overnight, blocked with 5% nonfat milk in PBS, and incubated with diluted DBS eluates (1:50 for anti-HlyE IgA, 1:100 for anti-LPS IgA, and 1:500 for anti-HlyE IgG and anti-LPS IgG, corresponding to final ELISA dilutions of 1:500, 1:1000, and 1:5000, respectively). Bound antibodies were detected with goat anti-human IgA and IgG antibodies conjugated with horseradish peroxidase (Jackson ImmunoResearch Laboratories Inc., PA, USA). The peroxidase activity on the chromogenic *O*-phenylenediamine dihydrochloride (OPD; Sigma-Aldrich Co., MO, USA) substrate was measured kinetically at regular intervals at 450 nm. The blank-adjusted sample readings on each plate were averaged, divided by the average blank-adjusted reading of the standards included on the plate (human chimeric monoclonal antibody for HlyE; plasma pool for LPS), multiplied by 100, and reported as ELISA units.

### Statistical Analysis

Comparisons between groups were made using Fisher’s exact test and Mann-Whitney U test, where appropriate. Receiver operating characteristic (ROC) and area under the curve (AUC) analyses were performed to assess the diagnostic performance of the assay. For multi-marker comparisons, we used logistic regression to model additive combinations of markers. The optimal cut-off values were calculated using Youden’s J statistic (Youden’s index). AUCs and their 95% confidence intervals (CIs) were estimated using the DeLong method. Sensitivity and specificity were estimated at the Youden index threshold, and their 95% CIs were estimated by stratified bootstrap (2000 resamples). Statistical significance was set at *p* < 0.05. Statistical analysis was performed in R version 4.3.3, using the *pROC* package.

## RESULTS

### Participant Characteristics

Of 459 children enrolled in the parent study, 426 underwent surgical intervention, and 424 had evaluable DBS-based anti-HlyE and anti-LPS IgA results (Fig 1). For this analysis, the children were classified as typhoid intestinal perforation (TIP) cases based on pathognomonic intraoperative findings (i.e., full-thickness perforation or pre-perforation lesions involving the antimesenteric border of the distal ileum, without an alternative cause) and as controls if they had a confirmed alternative etiology of acute abdomen (Table S1). Participants with suspected typhoid-related manifestations or possible missed TIP (e.g., cholecystitis, primary peritonitis, or intra-abdominal abscess without confirmed perforation) were excluded from subsequent analyses (*n* = 35). After exclusion, 389 children comprised the full cohort with 341 TIP cases and 48 controls. A subset of 154 children (123 TIP cases and 31 controls) were tested for anti-HlyE and anti-LPS IgA and IgG antibodies at baseline and convalescence (longitudinal sub-cohort).

**Fig 1:**
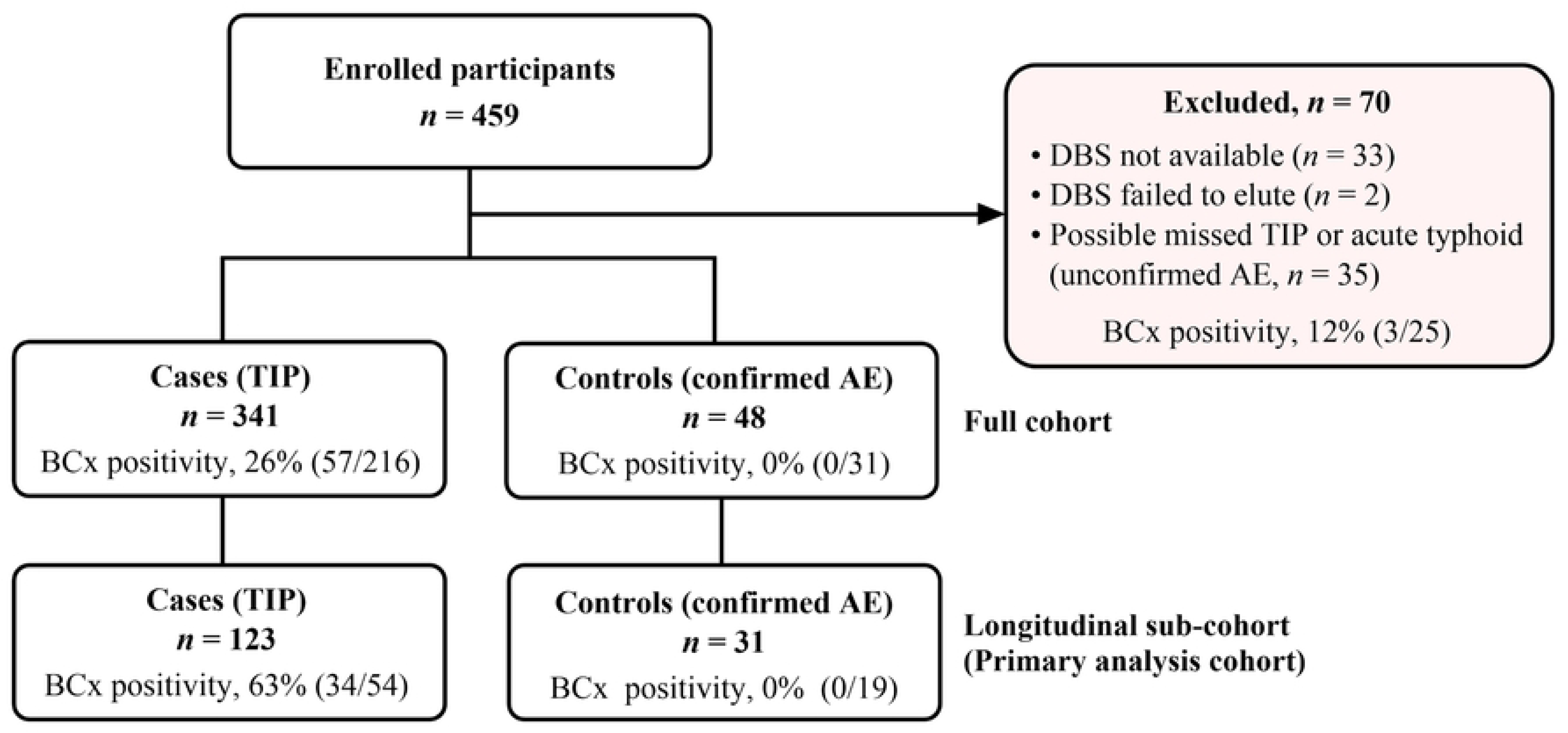
Classification of enrolled participants. BCx, blood culture; DBS, dried blood spot; TIP, typhoid intestinal perforation; AE, alternate etiology; possible typhoid-related disease without confirmed perforation (e.g., cholecystitis, unexplained peritonitis, intra-abdominal abscess).

In the full cohort, children with TIP were significantly younger than those in the control group (*p* < 0.0001), with an overall median age of 10 years (IQR 7–13) for TIP versus 13 years (IQR 10–15) for controls (Table 1). Fifty-eight percent of the cohort was male, and the distribution of sex was similar between cases and controls (*p* > 0.2736). Blood cultures were not available for all participants, as this procedure began after study enrollment. Of those with blood culture available, blood culture was positive for *S.* Typhi in 26% (57/216) of TIP cases, compared with 0% (0/31) of controls. *S.* Typhi was isolated from 12% (3/25) of blood cultures from children with suspected typhoid fever without confirmed TIP, lending support to their exclusion from analysis (Fig 1). The age and sex distribution of the sub-cohort was comparable to the full cohort (Table 1), but in TIP cases, blood culture positivity was higher in the longitudinal cohort (63%) compared to the full cohort (26%).

**Table 1:** Participant characteristics.

| Parameter | Control | TIP | Total | P-value |
| --- | --- | --- | --- | --- |
| **Full cohort** | **n = 48** | **n = 341** | **n = 389** |  |
| Age (years) | 13 (10–15) | 10 (7–13) | 10 (7–13) | < 0.0001 |
| < 10 years (%) | 7 (15) | 152 (45) | 159 (41) |  |
| ≥ 10 years (%) | 41 (85) | 189 (55) | 230 (59) |  |
| Female (%) | 16 (33) | 146 (43) | 162 (42) | 0.2736 |
| Male (%) | 32 (67) | 195 (57) | 227 (58) |  |
| Blood culture performed (%) | 31 (65) | 216 (63) | 247 (63) | > 0.9999 |
| *Salmonella* Typhi positive (%)* | 0 (0) | 57 (26) | 57 (23) | 0.0003 |
| **Longitudinal sub-cohort**<br>**(Primary analysis cohort)** | **n = 31** | **n = 123** | **n = 154** |  |
| Age (years) | 13 (12–15) | 10 (7–12) | 10 (8–13) | < 0.0001 |
| < 10 years (%) | 4 (13) | 60 (49) | 64 (42) |  |
| ≥ 10 years (%) | 27 (87) | 63 (51) | 90 (58) |  |
| Female (%) | 12 (39) | 48 (39) | 60 (39) | > 0.9999 |
| Male (%) | 19 (61) | 75 (61) | 94 (61) |  |
| Blood culture performed | 19 (61) | 54 (44) | 73 (47) | 0.1075 |
| *Salmonella* Typhi positive (%)* | 0 (0) | 34 (63) | 34 (47) | < 0.0001 |

Data are in n (%) and median (IQR). \*Proportions calculated based on the number of blood cultures performed. IQR, interquartile range; TIP, typhoid intestinal perforation.

### Distribution of Baseline Anti-HlyE and Anti-LPS IgG and IgA Responses

In the longitudinal sub-cohort, which included children with all four antigen-antibody measures, the median baseline level of each anti-HlyE- and anti-LPS-specific IgA and IgG antibody was significantly higher in the TIP cases than in controls (*p* < 0.0001; Fig 2A). Age-stratified analyses were limited by the small number of controls under 10 years of age (*n* = 4). Among children ≥ 10 years, TIP cases had significantly higher levels of all four biomarkers than controls (*p* < 0.0001; Fig 2B). Among children < 10 years of age, only anti-LPS IgA (*p* = 0.0388) and anti-LPS IgG (*p* = 0.0152) levels were significantly higher in TIP cases than in controls. Antibody responses among surgically diagnosed TIP cases did not vary between those with blood culture-confirmed TIP and those without blood culture confirmation (including those without a blood culture or those with blood cultures negative for *S.* Typhi) (Table S2).

**Fig 2:**
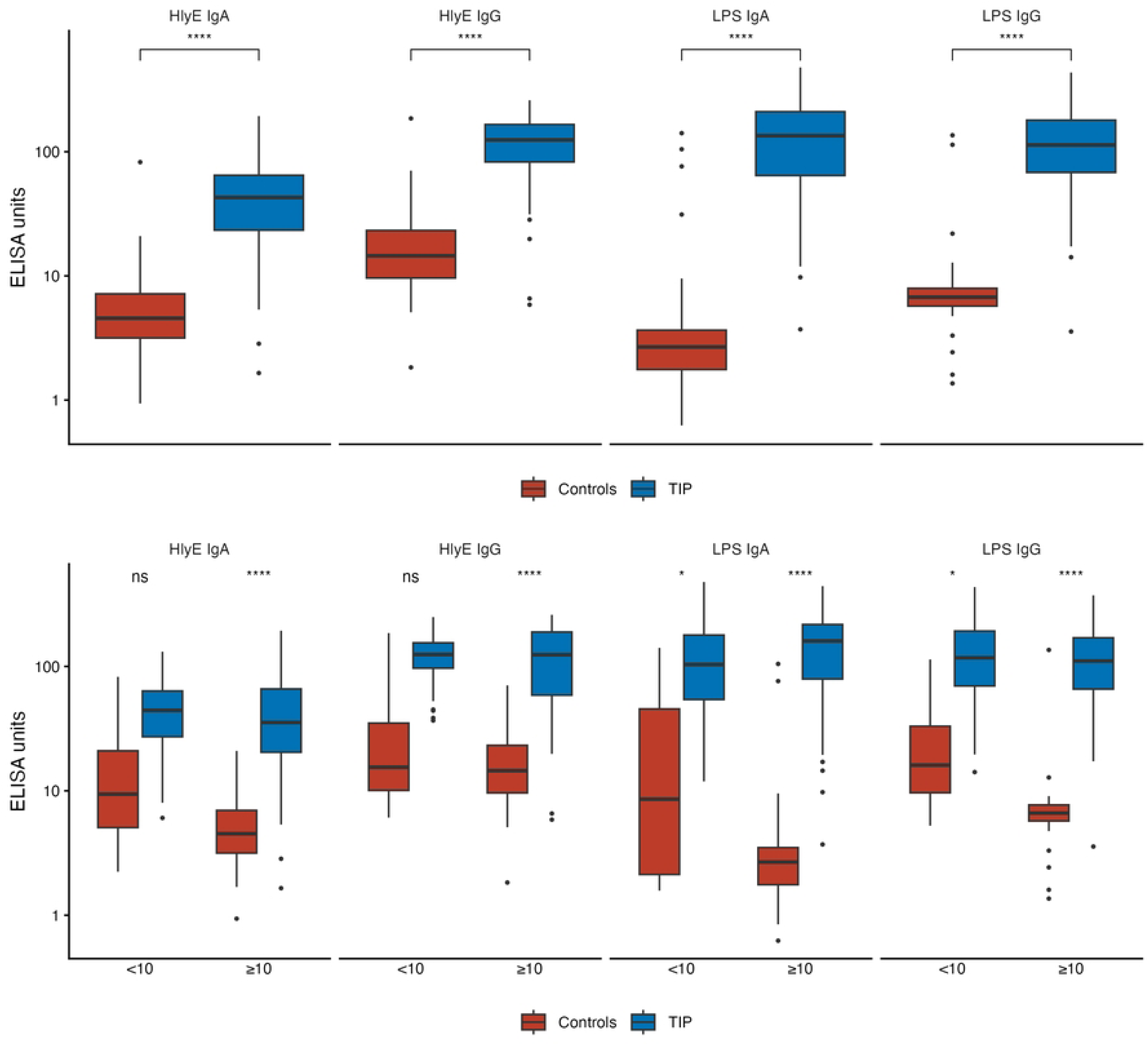
Distribution and comparison of anti-HlyE and anti-LPS IgA and IgG antibodies between cases and controls at baseline. Boxplots illustrate log (10)-transformed antibody titers among all participants (A) and stratified by age (years) in the longitudinal sub-cohort (B). *P*-values are between TIP cases and controls. BCx, blood culture; HlyE, hemolysin E; LPS, lipopolysaccharide; TIP, typhoid intestinal perforation. \**p* < 0.05, \*\*\*\**p* < 0.0001, ns = not significant (*p* > 0.05).

The anti-HlyE IgA and anti-LPS IgA results in the full cohort were consistent with those observed in the longitudinal sub-cohort, with TIP cases demonstrating higher anti-HlyE IgA and anti-LPS IgA levels than controls overall and among children ≥ 10 years of age (*p* < 0.0001, Fig S1A–B). In contrast to the longitudinal sub-cohort, both anti-HlyE IgA and anti-LPS IgA responses had significantly higher levels in TIP cases than controls among children < 10 years of age in the full cohort, likely reflecting the larger sample size in this age group (*p* ≤ 0.0221).

### Accuracy of anti-HlyE and anti-LPS for diagnosing TIP at presentation

The performance of each antigen and antibody isotype was assessed alone and in combination using the ROC AUC and Youden’s index to determine the best classification threshold within the primary analysis cohort, which had all measurements available. All evaluated individual antibodies and antibody combinations provided excellent discrimination between TIP cases and controls overall, with AUCs ≥ 0.94 and balanced accuracy ≥ 0.93 (Fig 3A; Table 2). Anti-LPS IgG demonstrated the highest discriminative ability among the individual markers (balanced accuracy, 0.96), with a sensitivity of 0.99 (95% CI: 0.94–1.00), and a specificity of 0.94 (95% CI: 0.84–1.00).

**Fig 3:**
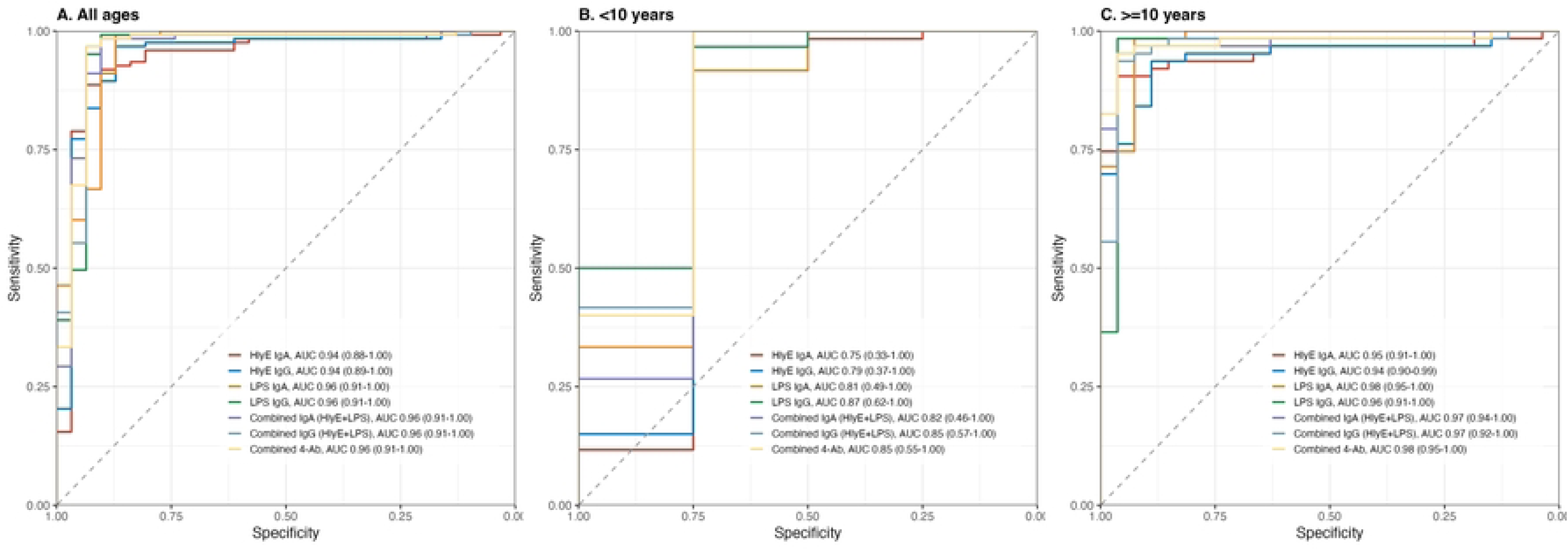
Receiver operating characteristic (ROC) area under the curve (AUC) for IgA and IgG responses to HlyE and LPS in the longitudinal sub-cohort at baseline. ROC curves are plotted for sensitivity versus specificity across all ages (**A**), < 10 years (**B**), and ≥ 10 years (**C**), and show the AUC (95% CI) values. CI, confidence interval; HlyE, hemolysin E; LPS, lipopolysaccharide; 4-Ab, all four antibodies.

**Table 2:** Diagnostic performance of anti-HlyE and anti-LPS IgA and IgG responses for TIP in the longitudinal sub-cohort at baseline.

| Model | AUC<br>(95% CI) | Sensitivity<br>(95% CI) | Specificity<br>(95% CI) | Balanced<br>accuracy<br>(95% CI) |
| --- | --- | --- | --- | --- |
| **All ages** |  |  |  |  |
| HlyE IgA | 0.94 (0.88–1.00) | 0.92 (0.83–0.98) | 0.94 (0.84–1.00) | 0.93 (0.87–0.97) |
| HlyE IgG | 0.94 (0.89–1.00) | 0.97 (0.82–0.99) | 0.90 (0.77–1.00) | 0.94 (0.87–0.97) |
| LPS IgA | 0.96 (0.91–1.00) | 0.99 (0.92–1.00) | 0.87 (0.74–0.97) | 0.93 (0.87–0.98) |
| LPS IgG | 0.96 (0.91–1.00) | 0.99 (0.94–1.00) | 0.94 (0.84–1.00) | 0.96 (0.90–1.00) |
| Combined IgA (HlyE+LPS) | 0.96 (0.91–1.00) | 0.98 (0.91–1.00) | 0.90 (0.81–1.00) | 0.94 (0.89–0.99) |
| Combined IgG (HlyE+LPS) | 0.96 (0.91–1.00) | 0.98 (0.94–1.00) | 0.94 (0.84–1.00) | 0.96 (0.90–0.99) |
| Combined 4-Ab | 0.96 (0.91–1.00) | 0.98 (0.94–1.00) | 0.94 (0.84–1.00) | 0.96 (0.91–1.00) |
| **< 10 years** |  |  |  |  |
| HlyE IgA | 0.75 (0.33–1.00) | 0.95 (0.83–1.00) | 0.75 (0.25–1.00) | 0.85 (0.62–1.00) |
| HlyE IgG | 0.79 (0.37–1.00) | 1.00 (1.00–1.00) | 0.75 (0.25–1.00) | 0.88 (0.63–1.00) |
| LPS IgA | 0.81 (0.50–1.00) | 0.93 (0.35–1.00) | 0.75 (0.50–1.00) | 0.84 (0.67–1.00) |
| LPS IgG | 0.87 (0.62–1.00) | 0.97 (0.47–1.00) | 0.75 (0.50–1.00) | 0.86 (0.73–1.00) |
| Combined IgA (HlyE+LPS) | 0.82 (0.46–1.00) | 1.00 (0.35–1.00) | 0.75 (0.50–1.00) | 0.88 (0.63–1.00) |
| Combined IgG (HlyE+LPS) | 0.85 (0.57–1.00) | 1.00 (0.43–1.00) | 0.75 (0.50–1.00) | 0.88 (0.72–1.00) |
| Combined 4-Ab | 0.85 (0.55–1.00) | 1.00 (0.38–1.00) | 0.75 (0.50–1.00) | 0.88 (0.69–1.00) |
| **≥ 10 years** |  |  |  |  |
| HlyE IgA | 0.95 (0.91–1.00) | 0.92 (0.83–0.97) | 0.96 (0.89–1.00) | 0.94 (0.88–0.98) |
| HlyE IgG | 0.95 (0.90–0.99) | 0.94 (0.78–0.98) | 0.93 (0.78–1.00) | 0.93 (0.85–0.97) |
| LPS IgA | 0.98 (0.95–1.00) | 0.98 (0.95–1.00) | 0.93 (0.82–1.00) | 0.96 (0.90–1.00) |
| LPS IgG | 0.96 (0.91–1.00) | 0.98 (0.95–1.00) | 0.96 (0.89–1.00) | 0.97 (0.93–1.00) |
| Combined IgA (HlyE+LPS) | 0.97 (0.94–1.00) | 0.97 (0.89–1.00) | 0.96 (0.89–1.00) | 0.97 (0.91–1.00) |
| Combined IgG (HlyE+LPS) | 0.97 (0.93–1.00) | 0.95 (0.89–1.00) | 0.96 (0.85–1.00) | 0.96 (0.91–0.99) |
| Combined 4-Ab | 0.98 (0.95–1.00) | 0.97 (0.87–1.00) | 0.96 (0.89–1.00) | 0.97 (0.92–1.00) |

AUC, area under the curve; CI, confidence interval; HlyE, hemolysin E; LPS, lipopolysaccharide; Combined 4-Ab, all four biomarkers.

To assess the performance of the antibody responses by age, we stratified the primary analysis cohort into two age strata: < 10 years and ≥ 10 years (Table 2). The < 10 years analysis was limited by the low number of controls in that age group (*n* = 4), with the performance of all antigen-antibody isotypes numerically lower, with AUCs ranging from 0.75 (95% CI: 0.33–1.00) for HlyE IgA to 0.87 (95% CI: 0.62– 1.00) for LPS IgG (Fig 3B), compared to the older age group. The discriminatory accuracy of the antibody responses in children ≥ 10 years was excellent for all antigen isotypes, with all AUCs ≥ 0.94 (sensitivities ranging from 0.92–0.98, and specificities ranging from 0.93–0.96 (Fig 3C). Anti-LPS IgG had the highest balanced accuracy of the individual markers (0.97), with a sensitivity and specificity of 0.98 (95% CI: 0.95–1.00) and 0.96 (95% CI: 0.89–1.00), respectively. The combination of anti-LPS IgA and anti-HlyE IgA performed comparably to anti-LPS IgG in this age group, with a sensitivity of 0.97 (95% CI: 0.89–1.00), and a specificity of 0.96 (95% CI: 0.89–1.00).

The full-cohort analysis confirmed the findings of the primary analytic cohort. Anti-HlyE IgA and anti-LPS IgA, either alone or combined, discriminated TIP cases from controls with excellent accuracy (AUCs 0.92–0.97) overall and among children ≥ 10 years of age; performance of these biomarkers remained lower in children < 10 years of age (Fig S2, Table S3).

### Convalescent Antibody Responses

Antibody responses were largely unchanged between presentation and convalescence within both TIP cases and controls, while the differences between groups remained significant at both time points (Fig S2). Accordingly, discrimination of TIP cases from controls was similar at presentation and convalescence, with no significant difference in ROC AUC estimates by DeLong’s test (Table S4).

## DISCUSSION

The anti-HlyE and anti-LPS antibodies have already shown promise as biomarkers for estimating typhoid incidence and diagnosing acute typhoid fever [6, 20, 25]. In this study, we demonstrate that these antibody responses also show excellent discriminatory performance between patients with TIP and surgical controls. Anti-LPS IgG emerged as the single marker with the best diagnostic performance (99% sensitivity and 94% specificity). However, the performance of all the individual antigen-antibody responses and their combinations was broadly comparable, with all yielding excellent discrimination of TIP cases from controls and similarly high sensitivity and specificity. These findings suggest that serological profiling may provide a valuable adjunct tool for identifying probable typhoid-associated intestinal perforation in settings where microbiologic confirmation is unavailable.

Accurate estimation of TIP burden remains challenging because diagnosis relies on intraoperative findings, cases are not routinely captured through systematic surveillance, and conventional diagnostic methods have limited sensitivity. In particular, blood cultures are often negative by the time intestinal perforation occurs because circulating bacterial loads may have declined and antibiotic exposure before hospitalization is common [26, 27]. Our findings of high antibody responses to HlyE and LPS when patients present with TIP are consistent with the biological expectation that antibody titers against *S.* Typhi antigens are likely elevated in the setting of severe or disseminated disease, especially when perforation represents a late manifestation of systemic infection. This suggests that serological responses can identify clinically relevant TIP cases and typhoid disease when microbiologic confirmation is lacking.

Serological assays for assessing typhoid disease burden and typhoid-related complications may help prioritize interventions, such as vaccination and water/sanitation measures. In Niger, TIP has been reported to account for approximately 35–69% of acute peritonitis cases, although estimates vary across regions, study populations, and age groups [13, 28, 29]. In the underlying cohort for this analysis, 81% of enrolled participants who were undergoing surgery for atraumatic acute abdomen had typhoid-related disease [22]. The burden of TIP appears to be especially high among children ≤ 15 years, who have accounted for a median of 56% of cases of TIP in Francophone Africa [30] and in Niger [29]. Mortality remains substantial, with reported rates varying across settings and ranging between 6–37% [5, 22, 29, 30], but up to 50% in adult women of childbearing age operated for TIP in Niger [13]. Accurate detection of typhoid fever and TIP is critical for guiding and evaluating typhoid conjugate vaccine (TCV) programs. Reliable estimates of disease burden and severe complications strengthen the evidence base for TCV use in high-burden regions, including Niger and other parts of SSA, where vaccination is both effective [31–33] and likely more cost-effective than treatment of advanced complications [28, 32]. Because currently available TCVs do not induce antibodies against HlyE or LPS, these serological markers may also provide valuable tools for tracking trends in severe typhoid disease following vaccine introduction and for complementing culture-based surveillance and mortality data.

In many low-resource settings where TIP is common, laboratory infrastructure for ELISA-based testing may not be available. Therefore, incorporating anti-LPS IgG measurements in rapid diagnostic tests (RDTs) and other point-of-care tests (POCTs) could support TIP diagnosis and surveillance of severe typhoid disease at district hospitals and at peripheral facilities where blood culture-based and ELISA-based diagnostics are not feasible. Most commercially available POCTs for typhoid fever primarily detect IgM responses to *S.* Typhi antigens. A smaller number of RDTs include IgG detection; however, acute typhoid fever diagnostic performance has generally been suboptimal [34]. An emerging POCT for enteric fever, the DPP Typhoid Assay, measures IgA responses to HlyE and LPS, with > 90% sensitivity and specificity for diagnosing enteric fever [18–20]. The combination of these IgA biomarkers also performed comparably to LPS IgG alone in diagnosing TIP in this study. However, the performance of the DPP Typhoid Assay for diagnosing TIP, in addition to other IgG-based LPS (O-antigen) rapid diagnostics, requires further evaluation. Alternatively, DBS samples could be collected and transported to regional laboratories for ELISA-based testing, as was done in this study. DBS collection does not require highly specialized training, sophisticated transport, cold-chain infrastructure, or complex elution equipment or procedures, making it a feasible sampling strategy in resource-limited settings [35, 36].

The primary impact of TIP serological diagnosis lies in surveillance and improving the estimates of the burden of typhoid-associated intestinal perforation rather than in guiding immediate clinical decision-making. Serologic diagnosis is unlikely to have a direct impact on acute TIP management, as all patients presenting with an acute abdomen require surgical evaluation and intervention regardless of the underlying etiology. However, serologic diagnosis can improve case ascertainment where surgical diagnosis may be inconclusive or delayed. Still, blood cultures should be obtained whenever feasible, as they provide essential information on antimicrobial susceptibility patterns that guide clinical management and inform surveillance of antimicrobial resistance.

Convalescent antibody measurements did not improve discrimination beyond the presentation (acute TIP) values, as similar antibody responses and AUC estimates were observed at both time points. The lack of a substantial rise in antibody levels between presentation and convalescence likely reflects the delayed timing of TIP relative to primary infection. Because perforation typically occurs after prolonged infection, patients may have already reached or approached peak humoral responses by the time of surgical presentation, limiting the additional diagnostic value of follow-up sampling [6, 37]. These findings suggest a single pre-operative measurement may be sufficient for diagnostic classification. This approach also offers practical advantages in resource-limited settings, where follow-up sampling is often difficult because of post-operative mortality, transportation barriers, and loss to follow-up, while also reducing testing costs [28].

The analysis has some limitations. First, the performance of these markers could not be fully evaluated in children under 10 years of age, given the low number of controls in this age group. Atraumatic intestinal perforation not due to typhoid fever is less common in this age group (i.e., appendicitis), and clinical presentation and surgical findings may differ from those in adults and older children. As a result, the generalizability of these findings to younger pediatric populations remains uncertain. In addition, the control population consisted of children with surgically confirmed alternative causes of acute abdomen rather than a broader population of febrile or hospitalized children. As a result, diagnostic performance may differ in other clinical settings and could be lower when evaluated against conditions with overlapping immunologic or clinical features. Second, the diagnosis of TIP was based primarily on characteristic intraoperative findings rather than microbiologic confirmation. Although these findings are widely used in clinical practice and have been shown to correlate strongly with typhoid etiology, some degree of outcome misclassification remains possible. Because serologic performance was evaluated against a clinical-surgical reference standard rather than a definitive microbiologic gold standard, estimates of diagnostic accuracy should be interpreted accordingly. We sought to minimize this possibility by excluding participants with clinical or surgical findings suggestive of possible TIP or severe typhoid fever and limiting controls to those with confirmed alternative etiologies. Lastly, the study was conducted at a single rural center, the Hôpital de la SIM-Galmi in Niger. Therefore, the findings may not be fully applicable to other settings.

In conclusion, these findings support the use of serological signatures to identify typhoid-associated intestinal perforation and distinguish it from other causes of acute abdominal disease in children. Such an approach could enhance surveillance of severe typhoid complications, improve burden estimates in endemic settings, and generate critical evidence to support TCV introduction and evaluation. Further validation, particularly in younger pediatric populations, is needed to refine and validate these findings.

## Data Availability

The URL link to the minimal data set will be provided at acceptance

## Acknowledgments

This study was supported by grants from the Gates Foundation (INV-042348, INV-042350, and INV-030857). The authors express their profound gratitude to all study participants, staff of the Department of Surgery of the Hôpital de la SIM-Galmi in Niger, and the Typhoid Vaccine Acceleration Consortium (TyVAC) for their valuable contributions to this study.

